# Inferring Alzheimer’s disease pathologic traits from clinical measures in living adults

**DOI:** 10.1101/2023.05.08.23289668

**Authors:** Jingjing Yang, Xizhu Liu, Shahram Oveisgharan, Andrea R. Zammit, Sukriti Nag, David A Bennett, Aron S Buchman

**Author notes:** **Corresponding Author:** Jingjing Yang, Address: Department of Human Genetics, Emory University School of Medicine, 615 Michael Street, Suite 305K, Atlanta, GA, 30322, USA.

## Abstract

**Background:** Alzheimer’s disease neuropathologic changes (AD-NC) are important for identify people with high risk for AD dementia (ADD) and subtyping ADD.

**Objective:** Develop imputation models based on clinical measures to infer AD-NC.

**Methods:** We used penalized generalized linear regression to train imputation models for four AD-NC traits (amyloid-*β*, tangles, global AD pathology, and pathologic AD) in Rush Memory and Aging Project decedents, using clinical measures at the last visit prior to death as predictors. We validated these models by inferring AD-NC traits with clinical measures at the last visit prior to death for independent Religious Orders Study (ROS) decedents. We inferred baseline AD-NC traits for all ROS participants at study entry, and then tested if inferred AD-NC traits at study entry predicted incident ADD and postmortem pathologic AD.

**Results:** Inferred AD-NC traits at the last visit prior to death were related to postmortem measures with *R^2^*=(0.188,0.316,0.262) respectively for amyloid-*β*, tangles, and global AD pathology, and prediction Area Under the receiver operating characteristic Curve (AUC) 0.765 for pathologic AD. Inferred baseline levels of all four AD-NC traits predicted ADD. The strongest prediction was obtained by the inferred baseline probabilities of pathologic AD with AUC=(0.919,0.896) for predicting the development of ADD in 3 and 5 years from baseline. The inferred baseline levels of all four AD-NC traits significantly discriminated pathologic AD profiled eight years later with p-values<1.4 × 10^−10^.

**Conclusion:** Inferred AD-NC traits based on clinical measures may provide effective AD biomarkers that can estimate the burden of AD-NC traits in aging adults.

## INTRODUCTION

The accumulation of Alzheimer’s disease neuropathologic changes (AD-NC), such as amyloid-*β* and intracellular neurofibrillary tangles, underlying Alzheimer’s disease dementia (ADD) has been observed even during the initial stages of ADD when cognition is normal [1]. Higher levels of AD-NC during the early stages of ADD have been shown to be associated with an increased risk of ADD [2–6]. Separate therapies have been developed for targeting the peptide amyloid-*β* in extracellular amyloid plaques and the protein tau in intracellular neurofibrillary tangles [7–9]. Yet, conventional prediction models identifying adults at risk for clinical ADD do not inform on which AD-NC traits underlie the risk of ADD. Thus, procedures that can accurately infer elevated levels of AD-NC traits in living adults with normal cognition have the potential to facilitate early targeted treatments [10–17].

Direct measures of brain AD-NC traits can only be obtained at autopsy. Recent efforts to quantify AD-NC during life have focused on identifying biomarkers of AD-NC traits by using brain imaging or fluid AD biomarkers [18–20]. Recent work comparing tau and amyloid positron emission tomography (PET) brain imaging measures to indices measured at autopsy, suggests that current imaging may not reliably detect the early stages of AD pathology [21, 22]. Particularly, brain imaging and CSF biomarkers are not widely available due to their costs, invasiveness, and difficultly to deploy at scale. Studies focusing on serum biomarkers have advanced rapidly in recent years, yet the field has not converged on specific biomarkers that can be employed in the general population [23–27].

Rapid advances in machine learning methods such as penalized generalized linear regression [28] have been employed to impute missing data or infer data that are difficult to be measured directly in biomedical research fields [29–34]. Similarly, the penalized generalized linear regression method could be deployed to develop imputation models based on clinical measures in older adults to infer levels of AD-NC traits. Such imputation models learn the predictive information of postmortem AD-NC traits like tangles from clinical measures obtained prior to death in decedents undergoing autopsy. The imputation model works by mathematically “explaining” the variation of observed AD-NC traits measured in decedents by their equivalence of weighted linear combinations of predictive clinical measures. Once imputation models developed for AD-NC traits are validated in an independent cohort, they can be applied to any older adults with similar clinical measures to infer AD-NC traits.

Comprehensive clinical and postmortem data are necessary to develop and validate imputation models for AD-NC traits. This multi-stage study leveraged clinical and postmortem data from two harmonized, independent, longitudinal prospective cohort studies –– Rush Memory and Aging Project (MAP) and the Religious Orders Study (ROS) [35]. First, we trained imputation models for four AD-NC traits (amyloid-*β*, tangles, global AD pathology, and pathologic AD diagnosis) by applying the penalized generalized linear regression method to clinical measures obtained at the last visit before death and postmortem AD-NC indices measured at autopsy in MAP decedents. Second, we validated the imputation models in a second independent cohort (ROS) which collected the same clinical and postmortem AD-NC traits. Third, we applied the imputation models to clinical measures collected in adults without dementia at study entry to infer baseline levels of AD-NC traits that were on average about eight years before death for decedents (or before last visit for living participants). We demonstrated the efficacy of these interfered baseline AD-NC traits as effective AD biomarkers, by showing their predictivity of future clinical ADD and discrimination of postmortem pathologic AD.

## MATERIALS AND METHODS

### Participants

Participants were community-dwelling older adults enrolled without known dementia and with at least two annual visits in one of two ongoing longitudinal prospective cohort studies of chronic conditions of aging –– MAP (n=1179 with ∼500 autopsied) and ROS (n=1103 with ∼600 autopsied). Both cohorts employed a harmonized data collection battery administered by the same research assistants facilitating joint analyses. For this study, we included adults without clinical evidence of dementia at enrollment with at least two annual follow-up cognitive assessments. At study entry, 1742 adults had no cognitive impairment (NCI) and 540 adults had mild cognitive impairment (MCI) (**Table S1**). The duration of annual follow-up for participants ranged from 2 to 26 years, with an average follow-up of 8 years (SD, 5.42 years) [**Fig S1; Tables S1-S2**].

### Assessment of AD-NC Traits

After the death of ROS/MAP participants, their brains were removed and hemisected following the standard procedure, as previously described [35]. Tissue blocks were dissected from predetermined regions and used for postmortem diagnosis of pathologic AD. Structured autopsy collected indices of AD/ADRD pathologies that were collected blinded to all prior clinical and cognitive data. This study focuses on the development of imputation models to infer the following four AD-NC traits that were measured in autopsied decedents (**Table S2**).

***Amyloid-β*** was labeled with an N-terminus–directed monoclonal antibody (10D5; Elan, Dublin, Ireland; 1:1,000). Immunohistochemistry was performed as previously described using diaminobenzidine as the reporter, with 2.5% nickel sulfate to enhance immunoreaction product contrast.

***PHFtau (Tangles)*** was labeled with an antibody specific for phosphorylated tau (AT8; Innogenetics, San Ramon, CA; 1:1,000). Amyloid-β load and tangles were quantified in 8 brain regions (anterior cingulate cortex, superior frontal cortex, mid frontal cortex, inferior temporal cortex, hippocampus, entorhinal cortex, angular gyrus/supramarginal gyrus, and calcarine cortex). Overall amyloid-β load was calculated through averaging mean percent area of amyloid-β deposition per region, across multiple brain regions. Tangles densities were derived by averaging tangles densities across corresponding brain regions. Measures of amyloid-*β* and tangles were further square-root transformed to improve their asymptotic normality as previously reported [36, 37].

***Global AD Pathology***: A modified Bielschowsky silver stain was used to visualize neuritic plaques, diffuse plaques, and neurofibrillary tangles in five cortical areas (hippocampus, entorhinal, midfrontal, middle temporal, and inferior parietal). Neuritic and diffuse plaques, and neurofibrillary tangles were counted in the region that appeared to have the maximum density of each pathology as previously described. A standardized score was created for each neuropathology in each region by dividing the raw count by the standard deviation of the mean for the same neuropathology in the same region. This standardization procedure puts the pathologic indices on a relatively common scale. A summary global AD pathology score was made based on the average of the greatest density of neuritic plaques, diffuse plaques, and neurofibrillary tangles in one mm^2^ [38, 39]

***Pathologic Diagnosis of AD***: The National Institute on Aging-Reagan criteria were used with intermediate and high likelihood cases indicating a pathologic diagnosis of AD, which is a binary indicator with value 1 denoting the present of pathologic AD and 0 denoting the absent of pathologic AD [40].

### Assessment of Composite Cognition Score and Cognitive Status

A structured cognitive assessment was administered annually. The neuropsychological battery included 19 tests that assessed five cognitive abilities (episodic memory, semantic memory, working memory, visuospatial ability/perceptual orientation, and perceptual speed). Raw test scores were standardized for each test using baseline means and standard deviations (SDs) of both cohorts; the resulting Z-scores were then averaged across these cognitive tests to derive a single summary composite cognition score as described in prior publications [35, 41].

Cognitive diagnoses were made in a three-step process. Cognitive testing was scored by a computer program and the results were reviewed by a neuropsychologist to diagnose cognitive impairment. Then participants were evaluated by a physician who used available cognitive and clinical data to classify cognitive status at each annual visit. Dementia required meaningful decline in cognitive function with impairment in multiple areas of cognition, and AD required dementia and progressive loss of episodic memory. Individuals with cognitive impairment who did not meet dementia criteria were diagnosed with mild cognitive impairment (MCI). Individuals without dementia or MCI were classified as having no cognitive impairment (NCI). Clinical diagnosis of cognitive status was based on published criteria [42–44]. Participants with dementia due to primary cause other than AD are excluded in this study. At the time of death, select clinical data from the entire study were reviewed by a neurologist, blinded to postmortem data, to render a final cognitive status diagnosis[35].

### Clinical Covariates

Diverse clinical measures were used to develop imputation models for four AD-NC traits. **Table S3** shows the complete list and groupings of the 57 clinical measures examined in this study, including both cross-sectional and longitudinal variables. Cross-sectional clinical measures were collected only once for each participant, such as sex, education, and *APOE* genotype. Longitudinal clinical measures were collected each time during participants’ annual visits. Baseline clinical characteristics are provided in **Tables S2, S4, and S5** and a heat map (**Fig S2**) is included to show the inter-correlations of the clinical variables examined in this study. The measures analyzed in this study were selected after excluding cross-sectional variables with a high proportion of missing values and those that were highly correlated with other selected clinical measures (correlation>0.95). Samples with missing values in the selected cross-sectional clinical measures were excluded. Missing values of longitudinal clinical measures were assumed to be missed at random and were imputed from the measured values at the nearest visit of the same participant, by using the “*fill(.direction = “up”)*” function from R library “*tidyr*”. That is, if a participant had a missing value at last visit for a longitudinal variable, the missing value would be imputed as the measured value of this variable in this participant’s nearest previous visit with collected measurement. The percentage of missing data (i.e., missing rate) in the longitudinal clinical measurements at last visit and study baseline were presented in **Fig, S3**. The number of samples that were actually used in the analyses are presented in **Tables S6**, and **S7**.

### Analytic Approach

A multi-stage analytic approach was employed to develop, validate, and demonstrate the effectiveness of the inferred the levels of four AD-NC traits at study entry as potential AD biomarkers.

### Developing imputation models to infer AD-NC traits

#### Stage 1. Training imputation models

We trained an imputation model for each of the four AD-NC traits using 57 clinical variables obtained in MAP participants at the last visit before death as predictors, by using the generalized linear regression model with Elastic-Net penalty (GLM-EN) [45] (**Fig 1A**). Only MAP decedents with autopsy were used for developing imputation models, because profiled AD-NC traits were required. By using the GLM-EN method, variable selection was implemented and potential collinearity among clinical variables was accounted for during model training. Since the Elastic-Net penalty is a linear combination of L1 (i.e. LASSO) [46] and L2 (i.e., Ridge) [47] penalties on the coefficients of clinical variables, variable selection are handled by the L1 penalty (i.e., penalizing the L1 norm of the coefficient vector) while potential collinearity is accounted for by the L2 penalty (i.e., penalizing the L2 norm of the coefficient vector). Ten-fold cross validation was used during model training to select Elastic-Net penalty parameters (i.e., the proportions of L1 and L2 penalty) to ensure optimal imputation accuracy.

**Fig 1.**
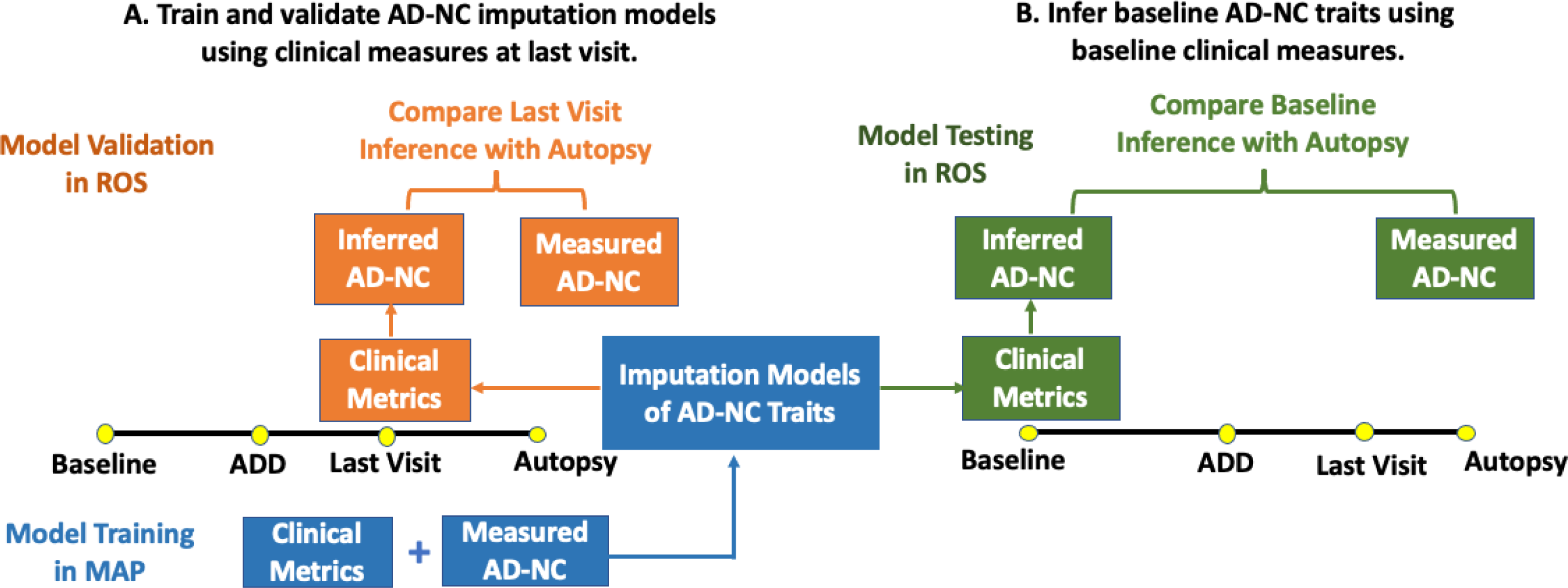
Overall study design to develop and validate imputation models that infer AD-NC traits based on clinical measures in older adults. A multi-stage analytic approach was employed to develop, validate, and demonstrate the effectiveness of inferred levels of four AD-NC traits derived from clinical measures as AD biomarkers. **A.** We trained imputation models for four AD-NC traits using clinical data obtained at the last visit before death in MAP decedents that underwent autopsy (**Fig 2**). Then we validated these models in an independent cohort study (ROS) that collected the same clinical and postmortem measures. **B**. We tested the effectiveness as AD biomarkers for the inferred levels of four AD-NC traits at baseline, which were obtained by applying the validated imputation models to clinical measures obtained at study entry. We examined if the inferred baseline AD-NC traits predicted incident ADD (**Fig 3**) and discriminated adults at risk for postmortem (on average 8 years after baseline) pathologic AD in ROS cohort (**Fig 4**).

#### Stage 2. Validating imputation models in a second independent cohort

Next, we validated the performance of the imputation models developed to infer four AD-NC traits using clinical variables measured at the last visit proximate to death in a second independent cohort with ROS decedents (**Fig 1A**). Prediction *R*^2^, the squared correlation between inferred and measured values, was used for assessing imputation accuracy for continuous AD-NC traits. Prediction accuracy for the dichotomous pathologic AD diagnosis was evaluated by the predicted area under curve (AUC) values of receiver operating characteristic curve (ROC) [48].

#### Stage 3. Infer AD-NC traits at study entry and test their effectiveness as AD biomarkers

Once imputation models are trained and validated, they can be applied to any living adult with the same clinical variables measured. So, to illustrate the use of these imputation models, we applied these validated models to the clinical data obtained from ROS participants at study entry (i.e., baseline that was on average of eight years before death or last follow-up visit) to infer baseline AD-NC traits (**Fig 1B**). Scatter plots and ROC curves were used to evaluate the consistency between inferred AD-NC traits at baseline and the corresponding measured postmortem AD-NC traits profiled at autopsy in ROS decedents. Then we examined the effectiveness of inferred AD-NC traits as potential AD biomarkers through two complementary analyses, one for evaluating the predictivity for incident ADD by Cox proportional hazard model (Stage 3A) and the other one for evaluating the discrimination of postmortem pathologic AD (Stage 3B).

Stage 3A. We fitted Cox proportional hazard models using covariates of age, sex, education, and a single inferred baseline AD-NC trait in MAP cohort for predicting incident ADD. Then we evaluated the performance of the Cox models for predicting incident ADD in year 3 and year 5 from baseline in ROS cohort (**Fig S4**). Our fitted Cox proportional hazard risk prediction models [49–51] also accounted for the competing risk of death. The annual cognitive status diagnosis and the follow up year were used to identify the first occurrence of ADD. For each participant, the year of enrollment is considered as baseline (time 0), the year of first diagnosis of ADD is considered as the time when the event occurs (incident ADD), and the last visit of participants without the considered event during all follow-ups is considered as the right censored time for living participants or the time of death for dead participants without ADD. Sample size distributions with respect to cognitive status at baseline and the cognition event types are shown in **Tables S1**. All Cox models were trained using data from MAP participants (both living and deceased) and tested with data from ROS participants (both living and deceased).

For fitting and testing Cox models for predicting incident ADD, we first used all individuals without dementia at baseline, and then fitted and tested another set of Cox models by using only individuals with NCI at baseline. For each set of Cox models fitted by using MAP participants, we calculated model accuracy (AUC) for predicting incident ADD in year 3 and year 5 from baseline.

Since Cox models provide a continuous risk score for incident ADD, by selecting a risk score threshold corresponding to ∼80% specificity (the proportion of correctly predicted non-ADD in test ROS participant’s, i.e., 1 – false positive fraction), we could calculate sensitivity (the proportion of true positive predictions in all test cases, i.e., true positive fraction), and the overall classification accuracy (the proportion of true discrimination of ADD in all test samples). Samples with risk scores greater than the selected threshold were considered to develop ADD and less than the threshold as not developing ADD in a specific year. That is, given the known ADD status of participants in year 3 and year 5 from baseline, we can compare the predicted risk of incident ADD to the actual incident ADD status to calculate the overall classification accuracy, specificity, and sensitivity. Also, the sensitivity and specificity corresponding to a selected risk score threshold reflect risk model performance at one point in the ROC plots.

We also sequentially added the other three inferred baseline AD-NC traits into the Cox model, in addition of covariates of age, sex, education, and inferred baseline amyloid-*β*, and examined the prediction performance of these models.

Stage 3B. We examined the discrimination of inferred baseline levels of AD-NC traits with respect to postmortem pathologic AD diagnosis (**Fig 1B**). Boxplots and two-sample t-tests were used to evaluate the discrimination of postmortem pathologic AD by inferred baseline AD-NC traits. Only ROS decedents with profiled pathologic AD diagnoses were use in this discrimination analysis.

## RESULTS

### Developed Imputation Models for Inferring AD-NC Traits

Imputation models were trained to infer AD-NC traits by applying the GLM-EN method using clinical measures in MAP decedents at their last visit before death as predictors. Selected predictive clinical measures with standardized effect sizes |beta| >0.01 estimated by the imputation models are shown in **Fig 2**. Composite cognition score and *APOE* E4 allele were the strongest predictors that were selected for all four AD-NC traits. Yet, **Fig 2** also illustrates that varied non-cognitive clinical measures including motor function such as motor gait and dexterity, health conditions such as anxiety and hypertension, and medications such as lipid lowering and anti-inflammatory medications were also selected. The effect size for motor gait was nearly as strong as APOE for tangles, amyloid-beta, and global AD pathology.

**Fig 2.**
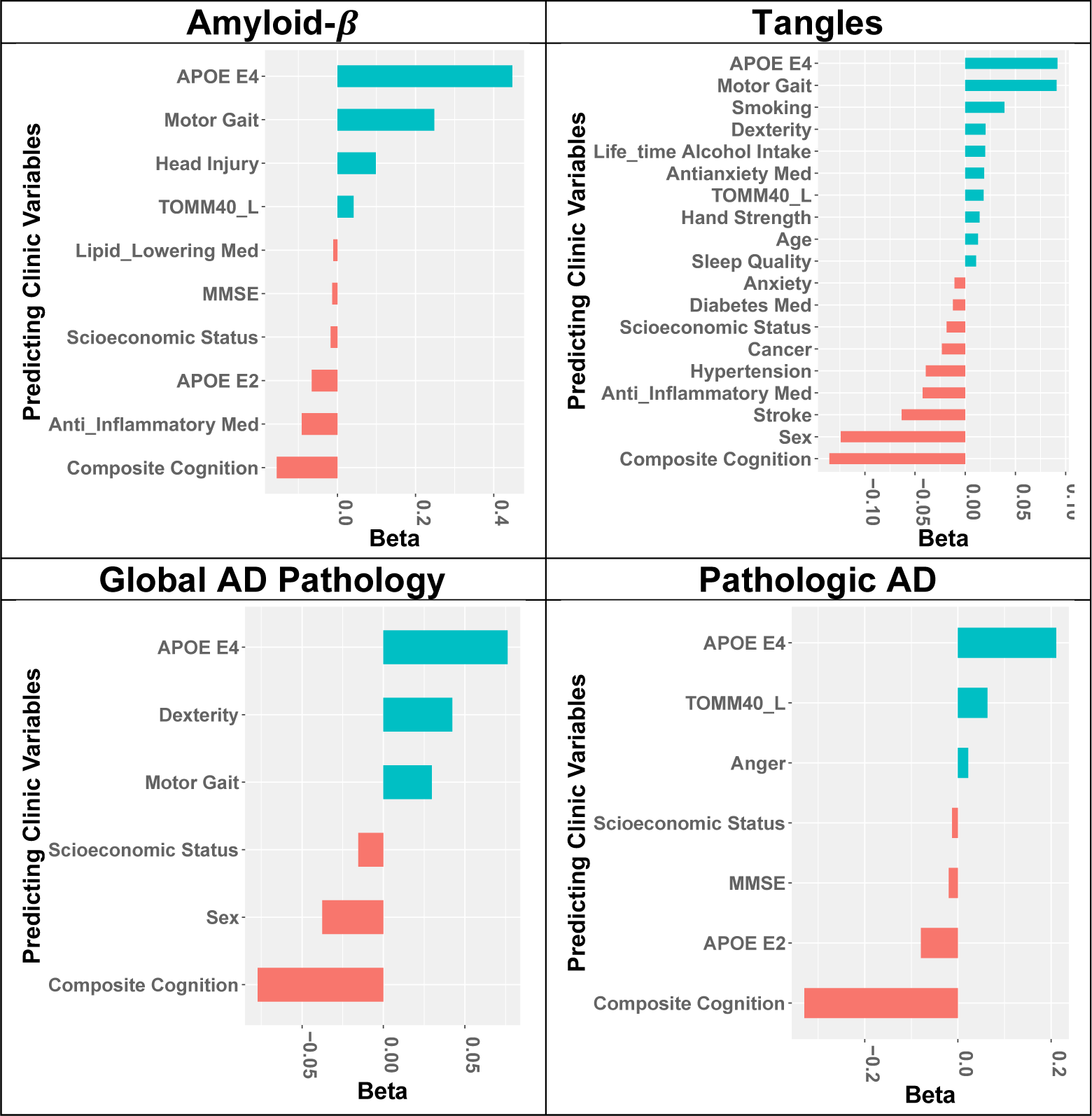
Machine learning methods were used to select different combinations of clinical measures to infer each of the four AD-NC traits. GLM-EN method was used to train an imputation model for each of the four AD-NC traits. Standardized effect sizes (beta) of selected predictive predictors with |beta| > 0.01 for each of the four imputation models were plotted. The inferred values of each of the inferred AD-NC traits are determined by the weighted averages of the corresponding selected predictors, with weights given by estimated standard effect sizes. Although all four AD-NC traits are inter-related and share cognition and *APOE E4* allele as important predictors, different sets of selected predictors by their imputation models highlight that different combinations of clinical measures with different effect sizes are necessary for inferring the unique features of these inter-related AD-NC traits.

These estimated effect sizes of selected clinical measure predictors in the imputation models are used as weights to construct weighted sum with clinical measurements to infer AD-NC trait levels. So, while all four AD-NC traits are related and may share composite cognition score and *APOE E4* allele as important predictors, the different sets of selected predictors by these four imputation models highlight that mathematically different combinations of clinical measures with different estimated effect sizes are necessary to best capture unique features of these inter-related AD-NC traits.

### Validation of Imputation Models for Inferring AD-NC Traits

To validate the imputation models developed in MAP decedents, we applied the imputation models to clinical measures obtained at the last visit prior to death for decedents in a second independent ROS cohort to infer their levels of four AD-NC traits. Scatter plots illustrate the correlations between the inferred levels of the three continuous AD-NC traits and their corresponding indices measured at autopsy (**Fig S5, A-C).** The prediction *R*^2^ was 0.188 for amyloid-*β*, 0.316 for tangles, and 0.262 for global AD pathology (**Table S8**). An ROC plot (with AUC=0.765) illustrates the consistency between the inferred probabilities of pathologic AD based on clinical measures obtained at last visit before death versus the profiled pathologic AD status by autopsy (**Fig S5D; Table S8)**. As would be expected for effective AD biomarkers, all four inferred AD-NC traits discriminated profiled pathologic AD at autopsy, with two-sample test P values < 10^−28^(Box plots in **Fig S5**; **Table S8**). Together these results in a second independent cohort validated the accuracy of the imputation models developed in MAP for all four AD-NC traits.

### Inferred Baseline AD-NC Traits Predicted Incident ADD

The imputation models were developed and validated using clinical measures at the last visit prior to death in MAP and ROS decedents. In further analyses we examined the effectiveness of inferred baseline AD-NC traits at study entry as AD biomarkers. To infer baseline levels of four AD-NC traits at study entry, we applied the validated imputation models to clinical data collected at baseline in all ROS participants (n=1103; both living and decedents), an average of 8 years before death for decedents (or last follow-up for living participants) (**Fig 1B**). Scatter plots of the three continuous AD-NC traits and an ROC plot of the binary pathologic AD illustrate the correlation between the inferred baseline versus the profiled AD-NC traits by autopsy (**Fig S6)**. Although the correlations were lower than the inferred AD-NC traits at last visit before death (**Fig S5**), the following analyses with Cox models still demonstrated the predictivity of the inferred baseline AD-NC traits for predicting incident ADD.

We employed separate Cox models that considered covariates age, sex, education, and each one of the four inferred baseline AD-NC traits to examine the predictivity for incident ADD (**Fig 3**). Coefficient estimates of the inferred baseline AD-NC traits in these Cox models were provided along with the corresponding p-values in **Table S9**. All inferred baseline AD-NC traits were strongly associated with incident ADD, with p-values < 10^−30^ in Cox models predicting incident ADD for adults without dementia at baseline, and p-values < 10^−5^ in Cox models for predicting incident ADD from adults with NCI at baseline.

**Fig 3.**
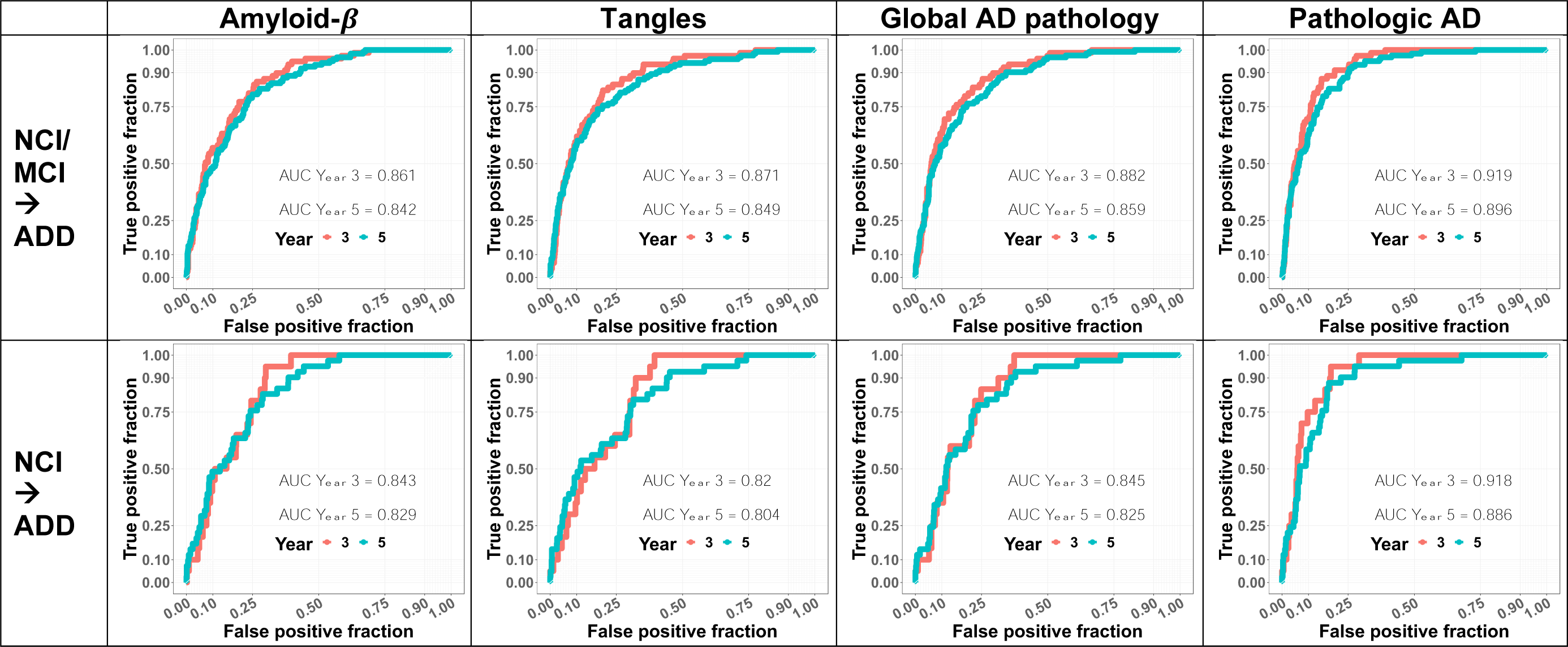
Inferred baseline AD-NC traits predicted incident Alzheimer’s Disease Dementia (ADD) We used Cox proportional hazard models to examine the predictivity of each of the inferred baseline AD-NC traits along with age, sex, and education covariates for incident ADD in 3 and 5 years after study entry. Top four panels show the prediction accuracies (ROC plots) with each the four inferred baseline AD-NC traits in adults without dementia (NCI+MCI) at study entry. Bottom four panels show prediction accuracies with each of the four AD-NC traits in adults with NCI at study entry. As expected for an effective AD biomarker, each of the inferred baseline AD-NC traits predicted ADD.

For all four AD-NC traits, model performance was higher in Year 3 (AUC ranging in 0.861 – 0.919) versus Year 5 (AUC ranging in 0.842 – 0.896) from baseline (**Fig 3, Upper Row**), for predicting incident ADD for adults without dementia at baseline. Of the four AD-NC traits, inferred baseline probabilities of pathologic AD had the highest predictivity (AUC 0.919 in Year 3; AUC 0.896 in Year 5) for incident ADD from adults without dementia at baseline. Similar results were observed when we restricted the analyses to the prediction of incident ADD in individuals with NCI at baseline (**Fig 3, Lower Row**).

By selecting a risk score threshold corresponding the ∼80% specificity, we calculated sensitivity and accuracy based on the prediction results for all of the Cox models (**Table 1**). Inferred baseline probabilities of pathologic AD also had the highest accuracy rates (80%) and sensitivity for predicting incident ADD in Year 3 (0.911) and Year 5 (0.829) from baseline (**Table 1**), compared to the other AD-NC traits.

**Table 1.** Prediction accuracy (with 95% confidence interval) and sensitivity with respect to selected risk score thresholds that ensure ∼80% specificity by Cox models using a single inferred AD-NC trait. Values in this table are reflecting the Cox risk model prediction performance at one point in the ROC curves as shown in **Fig 3**, with corresponding risk score thresholds. Samples with predicted risk scores greater than the selected threshold were considered as Predicted Positives (incident ADD), otherwise Predicted Negatives (not developing ADD).

| | | Amyloid-β | | Tangles | | Global AD Pathology | | Pathologic AD | |
| --- | --- | --- | --- | --- | --- | --- | --- | --- | --- |
|  |  | Y3 | Y5 | Y3 | Y5 | Y3 | Y5 | Y3 | Y5 |
| **NCI/MCI**<br>**->**<br>**ADD** | **Accuracy<sup>a</sup>**<br>**(95% CI)** | **0.798**<br>**(0.773,**<br>**0.822)** | **0.788**<br>**(0.763,**<br>**0.813)** | **0.802**<br>**(0.777,**<br>**0.825)** | **0.795**<br>**(0.771,**<br>**0.818)** | **0.801**<br>**(0.775,**<br>**0.823)** | **0.796**<br>**(0.771,**<br>**0.819)** | **0.809**<br>**(0.784,**<br>**0.831)** | **0.803**<br>**(0.778,**<br>**0.826)** |
|  | **Sensitivity<sup>b</sup>** | **0.772** | **0.699** | **0.822** | **0.756** | **0.797** | **0.764** | **0.911** | **0.829** |
|  | **Specificity<sup>c</sup>** | **0.800** | **0.800** | **0.800** | **0.800** | **0.800** | **0.800** | **0.800** | **0.800** |
| **NCI ->**<br>**ADD** | **Accuracy<sup>a</sup>**<br>**(95% CI)** | **0.796**<br>**(0.768,**<br>**0.823)** | **0.792**<br>**(0.763,**<br>**0.7819)** | **0.794**<br>**(0.765,**<br>**0.821)** | **0.791**<br>**(0.761,**<br>**0.818)** | **0.795**<br>**(0.767,**<br>**0.823)** | **0.793**<br>**(0.764,**<br>**0.820)** | **0.804**<br>**(0.775,**<br>**0.830)** | **0.805**<br>**(0.777,**<br>**0.832)** |
|  | **Sensitivity<sup>b</sup>** | **0.650** | **0.634** | **0.550** | **0.609** | **0.600** | **0.658** | **0.950** | **0.902** |
|  | **Specificity<sup>c</sup>** | **0.800** | **0.800** | **0.800** | **0.800** | **0.800** | **0.800** | **0.800** | **0.800** |

a. Accuracy= (# True Positive Predictions + # True Negative Predictions) / (# of test samples)
b. Sensitivity = (# True Positive Predictions) / (# Positives in test samples) = True Positive Fraction
c. Specificity = (# True Negative Predictions) / (# Negatives in test samples) = 1 – False Positive Fraction

In further analyses, we examined if adding covariates of additional inferred baseline AD-NC traits in a single Cox model would improve the prediction accuracy for incident ADD. As shown in **Fig S7 (Upper Row)**, we observed slightly improved AUC for prediction of incident ADD in adults without dementia, when we sequentially added each of the four AD-NC traits. Yet, modeling all four AD-NC traits together did not yield better results than using inferred probabilities of pathologic AD alone (**Last Column of Fig3 versus Fig S7**).

Sequentially adding the inferred baseline levels of three continuous AD-NC traits (amyloid-*β*, tangles, and global AD pathology) in a single Cox model did not improve the prediction of incident ADD for adults with baseline NCI (**Fig S7, Lower Row**). Although adding the inferred baseline levels of the probabilities of pathologic AD improved the prediction accuracy of incident ADD for adults with baseline NCI, but the prediction accuracy was comparable as using the inferred baseline levels of the probabilities of pathologic AD alone (**Last Column of Fig3 versus Fig S7**).

### Inferred Baseline AD-NC Traits Discriminated Postmortem Pathologic AD

By examining if the inferred baseline AD-NC traits would discriminate postmortem pathologic AD diagnosis, we presented boxplots of the inferred baseline AD-NC traits of ROS decedents with respect to their postmortem pathologic AD diagnosis by autopsy in **Fig 4**. By two-sample t-tests, we showed that all four inferred baseline AD-NC traits discriminated individuals with postmortem pathologic AD diagnosis by autopsy, with significant p-values < 1.4 x 10^−10^.

**Fig 4.**
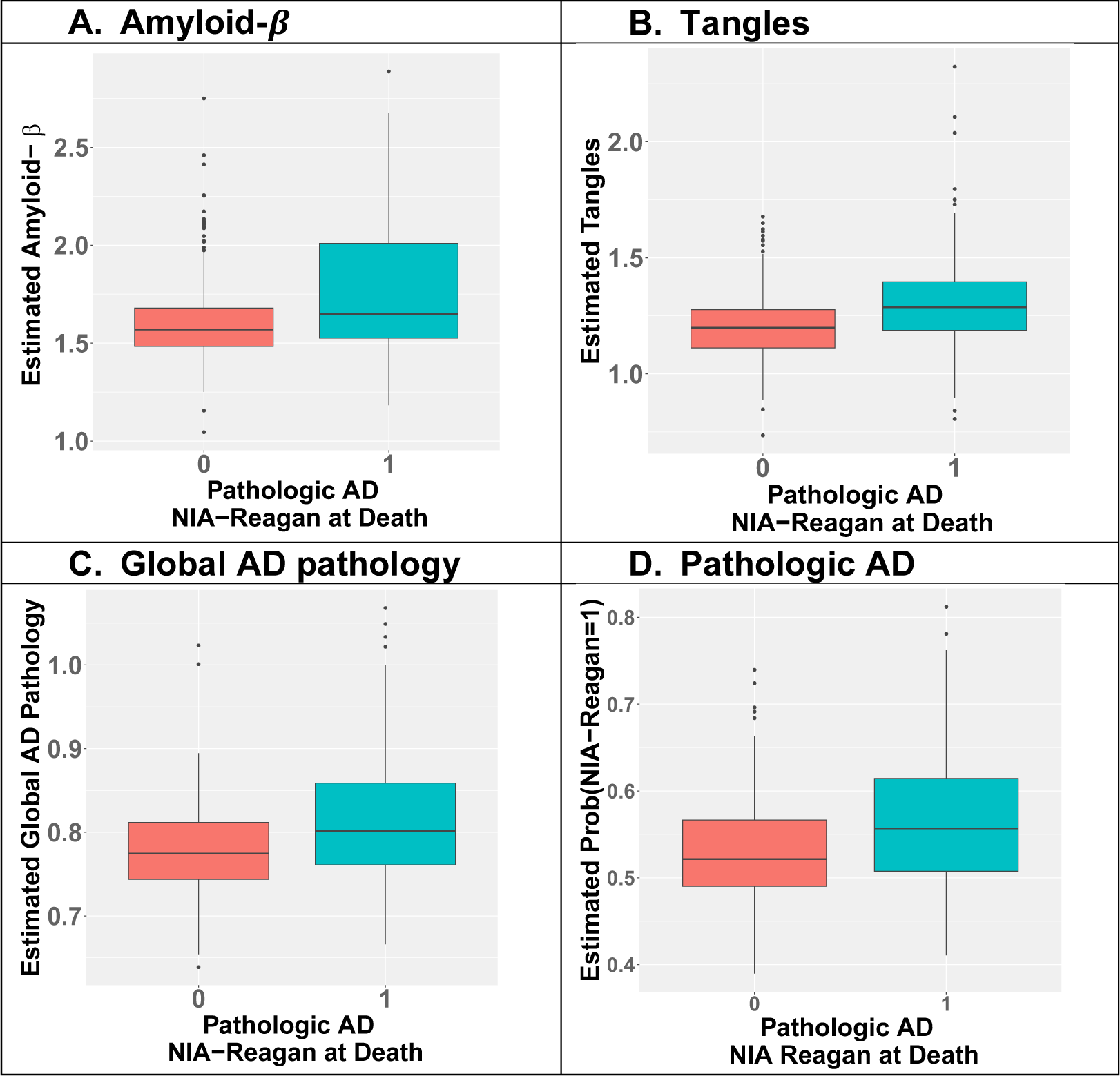
Inferred AD-NC traits at study baseline discriminated postmortem pathologic AD profiled at autopsy. Pathologic AD here is the binary postmortem NIA-Reagan status profiled at autopsy, with value 1 representing pathologic AD (teal boxplots) and 0 representing no pathologic AD (red boxplots). Two-sample t-test p-values are 1.4 × 10^−10^ for amyloid-β (A), 1.2 × 10^−11^ for tangles (B), 9.8 × 10^−12^ for global AD pathology (C), and 1.6 × 10^−10^ for pathologic AD (D).

## DISCUSSION

This study applied the machine learning GLM-EN methods to clinical measurements and postmortem indices of four AD-NC traits (amyloid-*β*, tangles, global AD pathology, and pathologic AD) obtained from the same older adults to develop imputation models that could be used to infer levels of four AD-NC traits based on clinical measurements. We validated these imputation models of AD-NC traits in a second independent cohort of older adults that collected similar clinical and postmortem indices. We applied the validated imputation models to clinical measures obtained at study entry to infer baseline AD-NC traits in adults about an average of eight years before death (decedents) or their last follow-up visit (living participants).

Adults without dementia at study entry who had higher baseline levels of inferred AD-NC traits had a higher risk of developing incident ADD during follow-up years and they also had a higher risk of having postmortem pathologic AD. These data suggest that inferred levels of AD-NC traits based on clinical measures may provide a low cost, non-invasive effective AD biomarker that can estimate the burden of AD-NC traits during the chronic course of Alzheimer’s disease. Inferred AD-NC traits may provide a way to monitor the clinical course of the accumulation of AD-NC traits underlying Alzheimer’s disease, improve the homogeneity of clinical trials and catalyze early targeted treatments to prevent the development of ADD in aging adults. Further studies to validate longitudinal inferred AD-NC traits will be needed.

### Novelty of this Study

Currently AD-NC traits in brain can only be measured at autopsy. Recent work has focused on identifying effective AD biomarkers that can be used to assess levels of AD-NC traits in living adults, especially during early stages of AD when cognition is still normal. There are many prior studies that have examined the associations of clinical measures, such as *APOE E4* allele, age, and cognitive measures, with future cognitive status or with different AD/ADRD indices measured at autopsy including the postmortem diagnosis of pathologic AD. Yet, it is important to note that the aim of these prior studies was not to infer levels of the different AD-NC traits examined in this study, nor to test their effectiveness as AD biomarkers [52–56].

Brain imaging studies of AD-NC traits have tried to employ serial imaging or CSF fluid biomarkers as proxies to obtain measures of AD-NC traits in brains to assess the accumulation of amyloid-*β* and tangles in early stages of Alzheimer’s disease [18–20]. Recent work that compared tau and amyloid PET brain imaging to AD indices measured at autopsy, suggests that current imaging may not reliably detect the early stages of AD pathology [21, 22]. Yet, the expense and limited availability of brain imaging and the invasiveness of obtaining CSF biomarkers make them difficult to be deployed at scale for the general population. This study fills this gap by employing machine learning methods that could be used to mathematically infer an AD-NC trait like “tangles” and estimate its burden at any time point prior to death in any adult with the requisite clinical measures.

Obtaining structured autopsy and diverse clinical measures during annual follow-ups, especially within a year prior of death, in large numbers of older adults is difficult. Thus, it is novel to have two large cohorts like MAP or ROS with the same clinical and postmortem indices of AD-NC traits that can be leveraged to develop imputation models in one cohort and validate these imputation models in a second independent sample of older adults. The rarity of these resources may explain in part the paucity of previous studies trying to infer AD-NC traits based on clinical measurements alone.

Another novel feature of this study is that we provide evidence that inferred baseline AD-NC traits on average 8 years before death for decedents (or before last visit for living participants) may be used as effective AD biomarkers as they predicted incident ADD and discriminated postmortem pathologic AD. That is, this study provides novel data demonstrating the feasibility and effectiveness of developing imputation models to infer AD-NC traits from clinical measures alone.

### Implications and Future Directions

This study is best conceptualized as an important first step highlighting that new machine learning analytic techniques can be used to infer AD-NC traits based on clinical measures collected in older adults. Further studies are still needed to determine if repeated inferred levels of AD-NC traits inform on trajectories of the accumulation of these different AD-NC traits. Currently, the temporal course of accumulation of these different AD-NC traits and the onset of their associations with impaired cognition are unknown. Such data are crucial to determine if inferred AD-NC traits could be used to assess the ongoing clinical course of Alzheimer’s disease, and to assess the efficacy of guiding treatments targeting specific AD-NC traits. For example, separate therapies have been under study for targeting the peptide amyloid-*β* in extracellular amyloid plaques and the protein tau in intracellular neurofibrillary tangles [7–9].

The analytic approach implemented in this study might also be extended to infer the presence of other pathologies that are hard to measure in living adults and untangle the effects of mixed-brain pathologies underlying late-life cognitive impairment and dementia. Since many older adults with ADD show mixed-brain pathologies [57], further work will be needed to develop analytic approaches that can infer and account for the presence of different combinations of varied AD/ADRD pathologies.

Brain imaging as well as serum or fluid biomarkers were not examined in this study, which could be used to validate the inferred levels of AD-NC traits by using our developed imputation models. Additionally, the brain imaging and fluid biomarkers might be included as additional predictors to enhance the imputation accuracy of AD-NC traits, which could be crucial for untangling the relative contributions of mixed-brain pathologies driving ADD in aging adults.

### Limitations and Strengths of this Study

This study still has several limitations. First, participants were predominantly Americans of European descent and have higher than average levels of education, so our findings will need to be replicated in more diverse populations. Second, the current study used diverse clinical predictors, many of that might not be available outside the research setting such as the composite cognition score based on 19 cognitive tests designed for ROS/MAP studies. Further work is needed to identify a parsimonious set of clinical predictors that are more widely available to enhance the use of this approach in diverse populations and geographic locations. Despite these limitations, this study is best conceptualized as an important first step highlighting the potential of using machine learning methods to infer AD-NC traits or other AD/ADRD pathologies based on clinical measures that can be collected via remote phenotyping or electronic health records.

Nonetheless, this study has several strengths that lend confidence for the current findings. All subjects were recruited from the community, underwent an annual detailed clinical evaluation, and were without dementia based on their clinical assessment at study entry. Large numbers of men and women underwent annual assessments, and follow-up rates were very high (∼90%) with an average of 8 years follow up. Uniform and structured procedures were employed for the collection of clinical measures and postmortem AD-NC traits in both MAP and ROS cohorts. An important strength of the current study design is that we developed imputation models in MAP cohorts, and then evaluated model performance in the second independent ROS cohort that employed similar staff and data collection procedures [58].

## ACKNOWLEGEMENTS

We are deeply indebted to all participants who contributed their data and agreed to autopsy at the time of their death. We thank the Rush Alzheimer’s Disease Center staff for their efforts.

## CONSENT STATEMENT

Both MAP and ROS studies were approved by an Institutional Review Board of Rush University Medical Center. All participants agreed to annual clinical evaluations and autopsy at the time of death. Written informed consent was obtained from all study participants as was an Anatomical Gift Act for organ donation.

## DATA AVAILABILITY

All data analyzed in this study are de-identified and available to any qualified investigator by submitting a request through the Rush Alzheimer’s Disease Center Research Resource Sharing Hub, https://www.radc.rush.edu, which has descriptions of the studies and available data.

