## Supplemental tables and figures for "Inferring Alzheimer’s disease pathologic traits from clinical measures in living adults"

#### Supplementary Tables

##### Table S1: Sample sizes for different outcomes.

| | Living Participants with NCI or MCI During all Follow-ups | Decedents without ADD | Decedents with ADD | Row Total |
|---|---|---|---|---|
| Baseline NCI | 976 | 397 | 369 | 1742 |
| Baseline MCI | 195 | 100 | 245 | 540 |
| Column Total | 1171 | 497 | 614 | 2282 |

##### Table S2. Clinical and postmortem characteristics of the analytic cohort (n=2,287).

| Group | Variable | Mean (SD) or N (%) |
|---|---|---|
| | Mild cognitive impairment | 541 (23.7) |
| | Follow-up years (2-26) | 8 (5.42) |
| i) Common AD risk factors | Age at baseline (years, 53-102) | 77.4 (7.51) |
| | Sex (males, N) | 577 (25.2) |
| | Education (years, 0-30) | 16.5 (3.65) |
| | MMSE Score (18-30) | 28.37 (1.73) |
| | APOE E2 (carriers, N) | 330 (14.4) |
| | APOE E4 (carriers, N) | 513 (22.4) |
| AD-NC traits profiled at autopsy | Amyloid-β | 1.73 (1.12) |
| | Tangles | 1.46 (0.47) |
| | Global AD Pathology | 0.84 (0.25) |
| | Pathologic AD | 771 (33.7) |

**Table S3. A list of clinical variables used to develop imputation models for AD pathology.**

| | Groups | Variables |
|---|---|---|
| i) | Common AD risk factors | Age, Sex, Education, MMSE Score, APOE Genotype E2 and E4 |
| ii) | Health conditions | BMI, Blood pressure (diastolic, hypertension), Depression score, Hypertension, Cancer, Diabetes, Head injury, Thyroid disease, Claudication, Heart disease, Stroke, Cardiovascular disease, Alcohol usage (in the past year and when drank most in lifetime), Smoking, |
| iii) | Medications | Usage of mental health, Analgesic, Antibiotic, Anti-hypertensive, Cardiac, Anti-anxiety, Anti-inflammatory, Aspirin, Lipid lowering, Insomnia, Diabetes medications |
| iv) | Clinical variables uniquely profiled by ROS/MAP | Global cognition test score, Physical activity, Social activity, Maternal and paternal education, Anxiety, Neuroticism indicating, proneness to psychological distress, Early life socioeconomic status, Anger trait, TOMM40 haplotype (S, L, VL) |
| v) | Motor and sleep metrics | Gait speed, Motor function, Dexterity, Gait, Hand strength, Bradykinesia score, Gait score, Global parkinsonian score, Rigidity score, Tremor score, 4 survey questions about sleeping status |

**Table S4. Baseline characteristics of clinical variables of groups (ii) and (iii) in Table S3, for the analytic cohort.**

| Group | Variable | Mean (SD) or N (%) |
|---|---|---|
| ii) Health conditions | BMI (range, 9.09 – 62.91) | 27.52 (5.50) |
| | Diastolic blood pressure (range, 40 – 122.5) | 74.47 (11.90) |
| | Hypertension blood pressure (range, 83 – 215.5) | 134.13 (18.29) |
| | Depression score (0 – 9) | 0.94 (1.48) |
| | Hypertension | 1142 (49.93) |
| | Cancer | 719 (31.43) |
| | Diabetes | 277 (12.11) |
| | Head injury | 148 (6.47) |
| | Thyroid disease | 440 (19.23) |
| | Claudication | 138 (6.03) |
| | Heart disease | 208 (9.09) |
| | Stroke | 156 (6.82) |
| | Cardiovascular disease history counts (0 – 3) | 0.93 (0.78) |
| | Alcohol usage in the past year, grams per day (0 – 116.55) | 4.56 (11.14) |
| | Alcohol usage when drank most in lifetime (0 – 6) | 0.46 (0.98) |
| | Smoking (never smoked 0, former smoker 1, current smoker 2) | 0.33 (0.51) |
| iii) Medications | Mental health | 529 (23.13) |
| | Analgesic | 1677 (73.32) |
| | Antibiotic | 158 (6.90) |
| | Anti-hypertensive | 1394 (60.95) |
| | Cardiac | 228 (9.96) |
| | Anti-anxiety | 133 (5.81) |
| | Anti-inflammatory | 561 (24.52) |
| | Aspirin | 992 (43.37) |
| | Lipid lowering | 740 (32.35) |
| | Insomnia | 171 (7.47) |
| | Diabetes | 203 (8.87) |

**Table S5. Baseline characteristics of clinical variables of groups (iv) and (v), for the whole analytic cohort.**

| | Variable | Mean (SD) or N (%) |
|---|---|---|
| iv) Clinical variables uniquely profiled by ROS/MAP | Global cognition test Z-score (-1.95 – 1.47) | 0.32 (0.51) |
| | Physical activity hours (0 – 35) | 3.19 (3.79) |
| | Social activity (0 – 162) | 7.58 (7.52) |
| | Maternal education (years, 0 – 20) | 9.66 (3.79) |
| | Paternal education (years, 0 – 24) | 9.60 (4.26) |
| | Anxiety Score (0 – 10) | 1.28 (1.69) |
| | Proneness to psychological distress (score, 0 – 42) | 15.69 (6.44) |
| | Tendency to be social, active, and optimistic (score, 5 – 24) | 15.52 (3.14) |
| | Early life socioeconomic status Z-score (-2.72 – 2.15) | 0.02 (0.74) |
| | Anger (score, 0 – 10) | 1.76 (1.88) |
| | TOMM40_S | 1,126 (49.2) |
| | TOMM40_L | 371 (16.2) |
| | TOMM40_VL | 1,114 (48.7) |
| v) Motor and sleep metrics | Gait speed (range, 0.05 – 1.57) | 0.63 (0.21) |
| | Motor function (score, 0.21 – 1.81) | 1.03 (0.23) |
| | Motor Dexterity (score, 0 – 1.85) | 1.01 (0.18) |
| | Motor gait (score, 0 – 2.13) | 1.02 (0.27) |
| | Motor hand strength (score, 0 – 2.36) | 1.03 (0.31) |
| | Bradykinesia score (0 – 85) | 9.45 (11.14) |
| | Gait score (0 – 85.18) | 12.16 (13.45) |
| | Global parkinsonian score (0 – 52.30) | 6.33 (6.30) |
| | Rigidity score (0 – 60) | 2.15 (5.88) |
| | Tremor score (0 – 69.69) | 2.23 (5.02) |
| | Sleep Latency (score, 1 – 9) | 3.63 (1.14) |
| | Sleep Consolidation (score, 1 – 8) | 2.92 (1.31) |
| | Daytime Sleepiness (score, 1 – 5) | 3.21 (1.33) |
| | Sleep Quality (score, 1– 5) | 2.10 (1.05) |

**Table S6: Sample sizes (the number of decedents) used for training and test imputation models for AD-NC traits.** Sample size differences are mainly due to the variation of the number of decedents whose corresponding AD-NC traits were measured after death.

| | Training Sample Size in MAP | Test Sample Size in ROS |
|---|---|---|
| ***Amyloid-β*** | 496 | 605 |
| ***Tangles*** | 509 | 605 |
| ***Global AD Pathology*** | 521 | 628 |
| ***Pathologic AD*** | 521 | 630 |

**Table S7: Sample sizes (both living and deceased participants with inferred AD-NC traits at baseline) used for training and test the Cox proportional hazard models as shown in Fig 3.**

| Cox Models | Training Sample Size in MAP | Test Sample Size in ROS |
|---|---|---|
| Adults with NCI or MCI at Baseline | 1183 | 1104 |
| Only Adults with NCI at Baseline | 905 | 837 |

**Table S8. Imputation model validation results in ROS decedents for AD-NC traits.** P-values were obtained for comparing inferred AD-NC traits at last visit to the corresponding postmortem measurements, by correlation test for continuous AD-NC traits including amyloid-$\beta$, tangles, global AD pathology, and by two-sample t-test for the binary pathologic AD diagnosis.

| | ***Amyloid-β ($R^2$)*** | ***Tangles ($R^2$)*** | ***Global AD Pathology ($R^2$)*** | ***Pathologic AD (ROC/AUC)*** |
|---|---|---|---|---|
| ***$R^2$ or ROC/AUC*** | **0.188** | **0.316** | **0.262** | **0.765** |
| ***P-value*** | $\mathbf{3.98 \times 10^{-29}}$ | $\mathbf{8.78 \times 10^{-52}}$ | $\mathbf{3.19 \times 10^{-43}}$ | $\mathbf{1.22 \times 10^{-33}}$ |

**Table S9.** ***Coefficient estimates of inferred baseline AD-NC traits and the corresponding p-values in the Cox proportional hazard models that were trained in MAP samples.*** *Four covariates of "age at baseline + sex + education + single inferred baseline AD-NC trait" were considered in the respective Cox model, where the inferred baseline AD-NC traits were standardized so that all coefficients were comparable.*

| **Inferred AD-NC traits** | **Coefficient estimate** | | **P-value** | |
|---|---|---|---|---|
| | **NCI/MCI → ADD** | **NCI → ADD** | **NCI/MCI → ADD** | **NCI → ADD** |
| ***Amyloid-β*** | **0.712** | **0.531** | $\mathbf{1.29 \times 10^{-31}}$ | $\mathbf{4.66 \times 10^{-9}}$ |
| ***Tangles*** | **0.982** | **0.617** | $\mathbf{2.07 \times 10^{-32}}$ | $\mathbf{2.91 \times 10^{-6}}$ |
| ***Global AD Pathology*** | **0.964** | **0.787** | $\mathbf{6.92 \times 10^{-45}}$ | $\mathbf{3.93 \times 10^{-12}}$ |
| ***Pathologic AD*** | **1.351** | **1.176** | $\mathbf{1.59 \times 10^{-58}}$ | $\mathbf{5.41 \times 10^{-20}}$ |

### Supplementary Figures

***Fig S1. Participants used to develop and validate imputation models for AD-NC.***
*Histogram plots of follow-up years (X-axis) and number of decedents (Y-axis) that were used to fit imputation models for AD-NC traits (A); follow-up years (X-axis) and  the number of living and deceased participants for assessing the predictivity for ADD at study entry (B).*

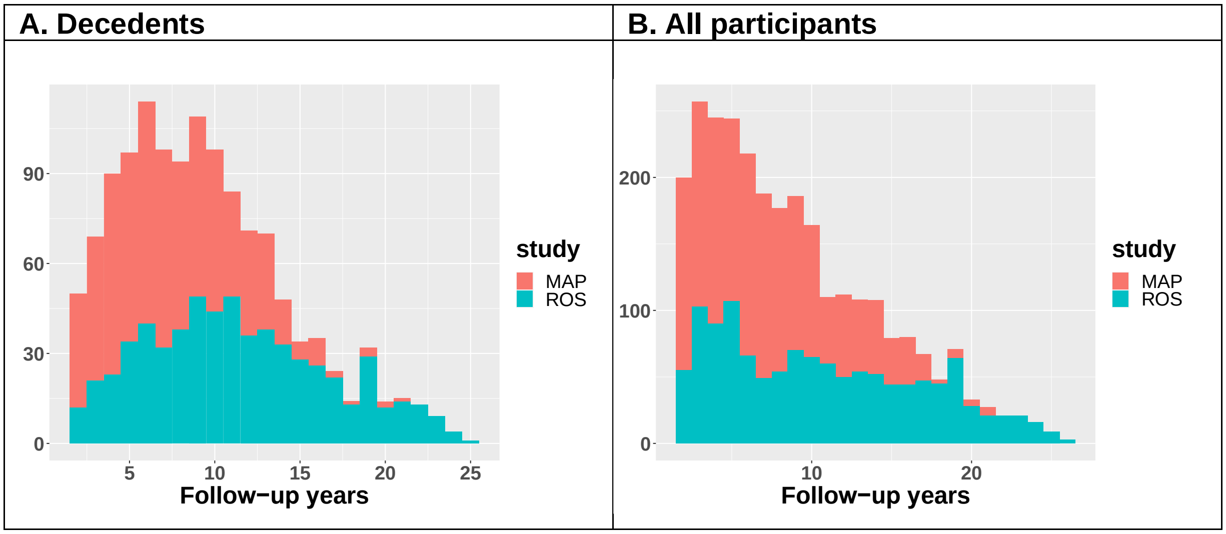

**Fig S2. Correlation heatmap of clinical variables used to develop imputations models for AD-NC traits.**

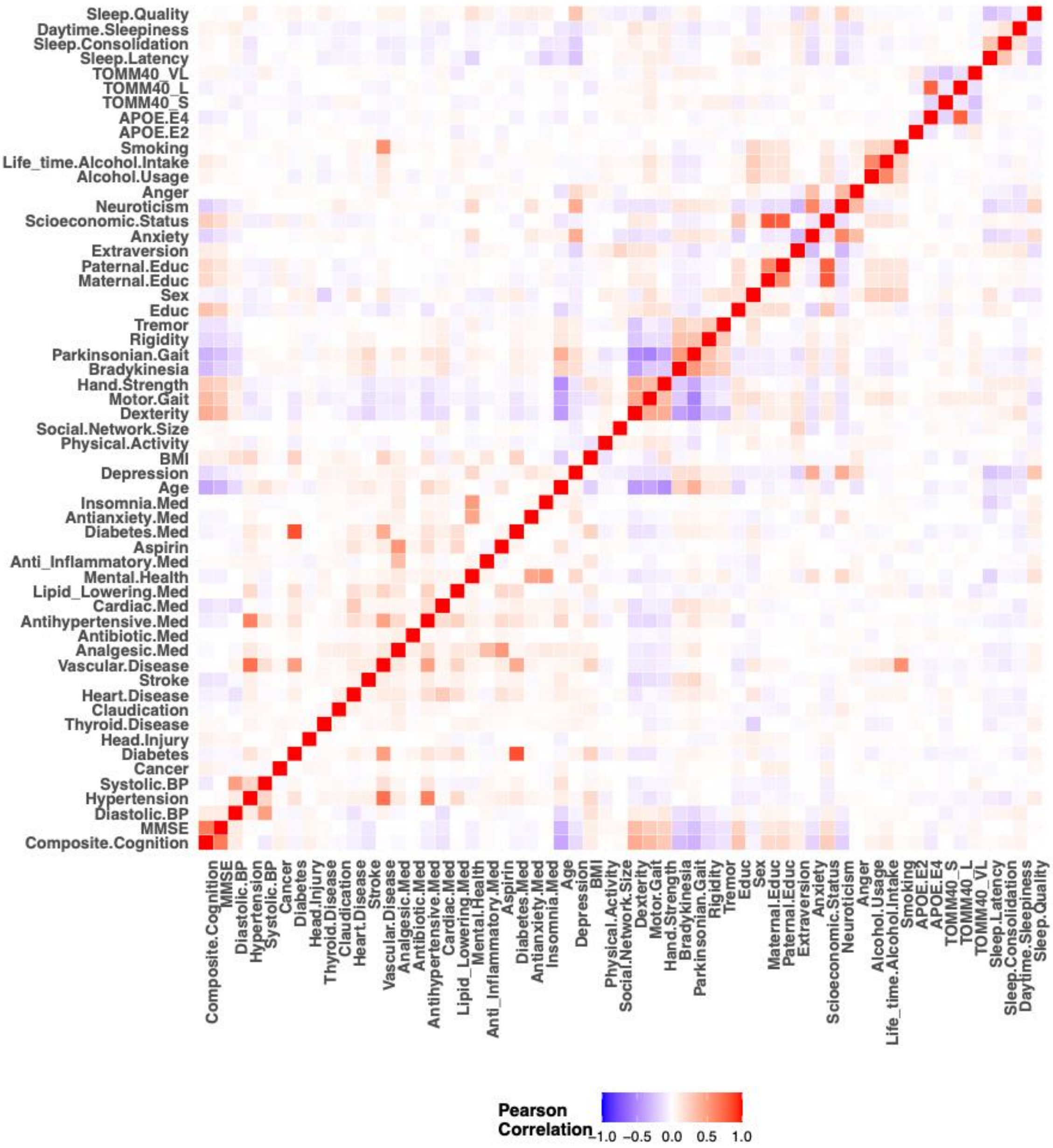

**Fig S3. Bar plots of the missing rates of clinical measures used to develop imputation models at last visit (A) and to infer AD-NC traits at study entry (B).**

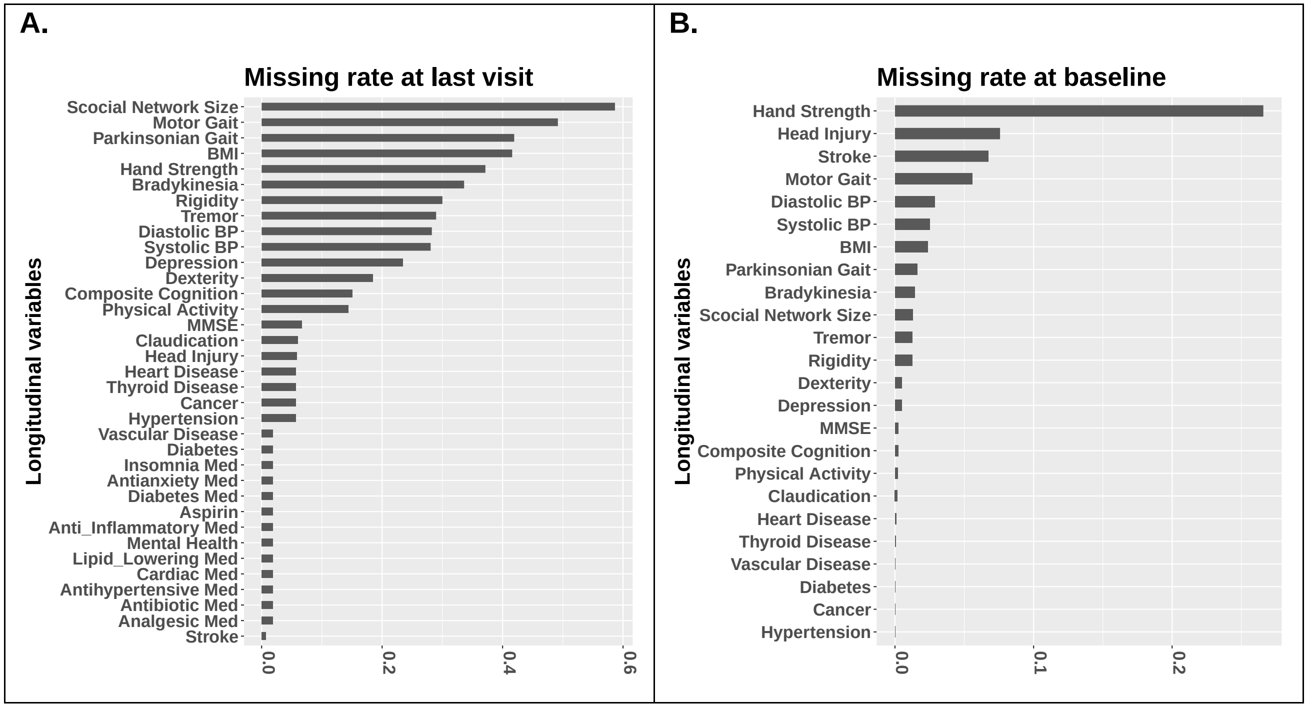

**Fig S4. Flowchart of evaluating the predictivity of inferred AD-NC traits at study entry for incident ADD.**

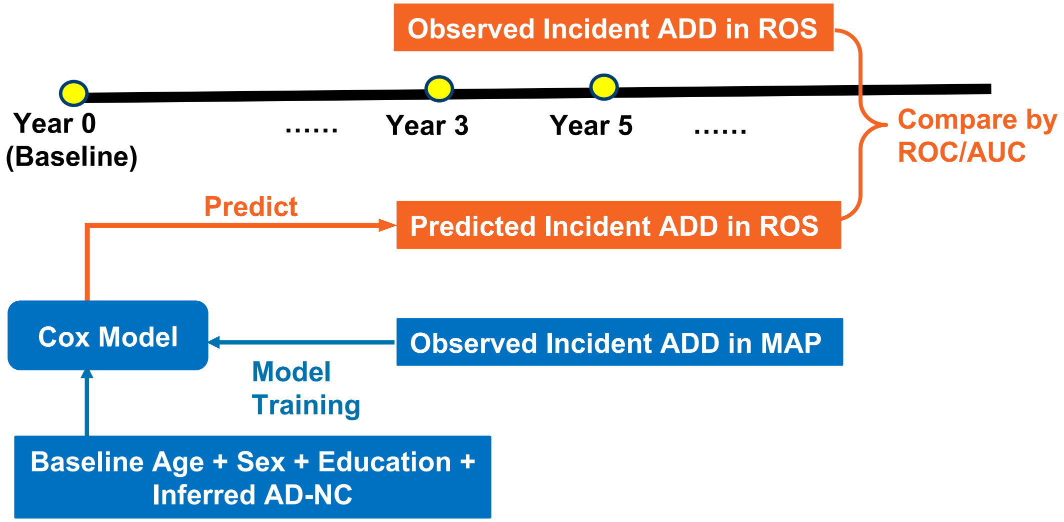

**Fig S5. Scatter plots and ROC comparing inferred AD-NC traits at last visit (proximate to death) in ROS decedents versus the corresponding postmortem measurements, and box plots showing the discriminations by inferred AD-NC traits with respect to postmortem pathologic AD.** Pathologic AD here is the binary NIA-Reagan status profiled at autopsy, with value 1 representing pathologic AD (teal) and 0 representing no pathologic AD (red). Two sample t-test p-values for testing inferred AD-NC traits versus postmortem pathologic AD are $1.07 \times 10^{-26}$ for amyloid-$\beta$ (**A**), $1.956 \times 10^{-30}$ for tangles (**B**); $1.47 \times 10^{-30}$ for global AD pathology (**C**); $1.22 \times 10^{-33}$ for pathologic AD (**D**).

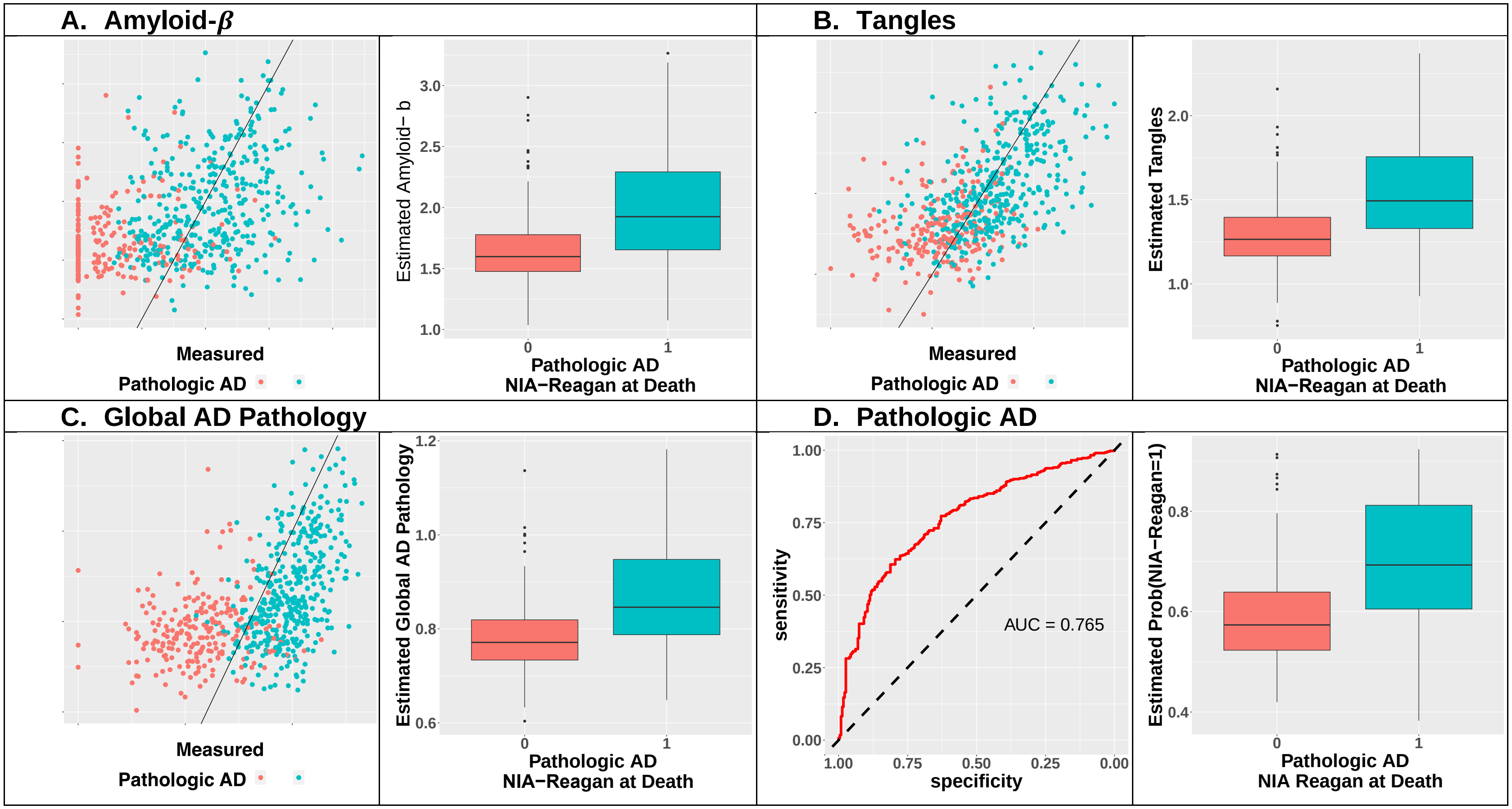

**Fig S6. Scatter plots and ROC comparing inferred AD-NC traits at baseline in ROS decedents versus the corresponding postmortem measurements.** Scatter plots for continuous AD-NC traits (**A, B, C**) and ROC plot for the binary pathologic AD (**D**) show that the inferred AD-NC traits based on clinical measures at study entry were correlated with their corresponding measures profiled at the time of autopsy about eight years later. Pathologic AD here is the binary NIA-Reagan status at autopsy, with value 1 representing pathologic AD (teal dots) and 0 representing no pathologic AD (red dots).

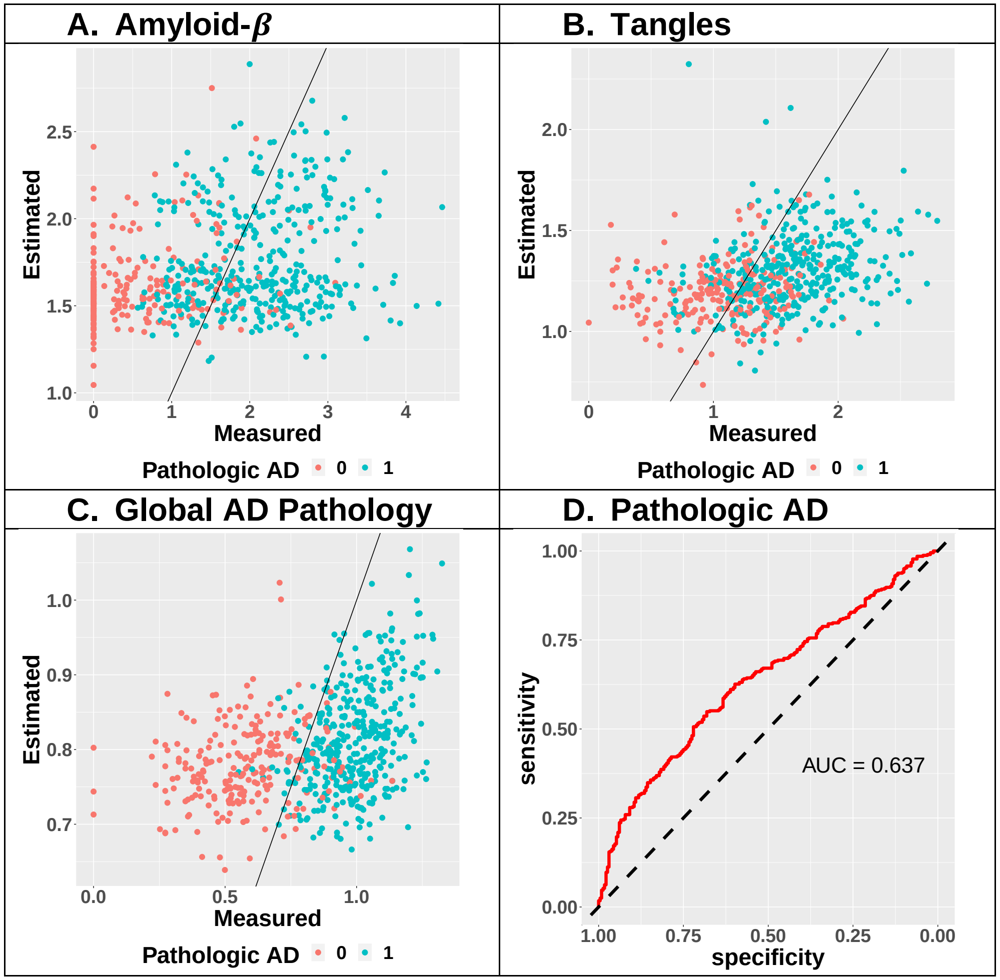

**Fig S7. Risk prediction results of ADD in year 3 and 5 after study entry for all ROS samples, by Cox proportional hazard regression models trained with MAP samples, using only baseline data of: age + sex + education + sequentially added inferred AD-NC traits at study entry.** Row 1 uses all samples without ADD (either NCI or MCI) at baseline, and row 2 uses only samples with NCI at baseline. By sequentially adding imputed pathologic AD to the other three AD pathology indexes, risk prediction accuracy is significantly improved, but is comparable as using inferred baseline pathologic AD alone as shown in the last column in Figure 3.

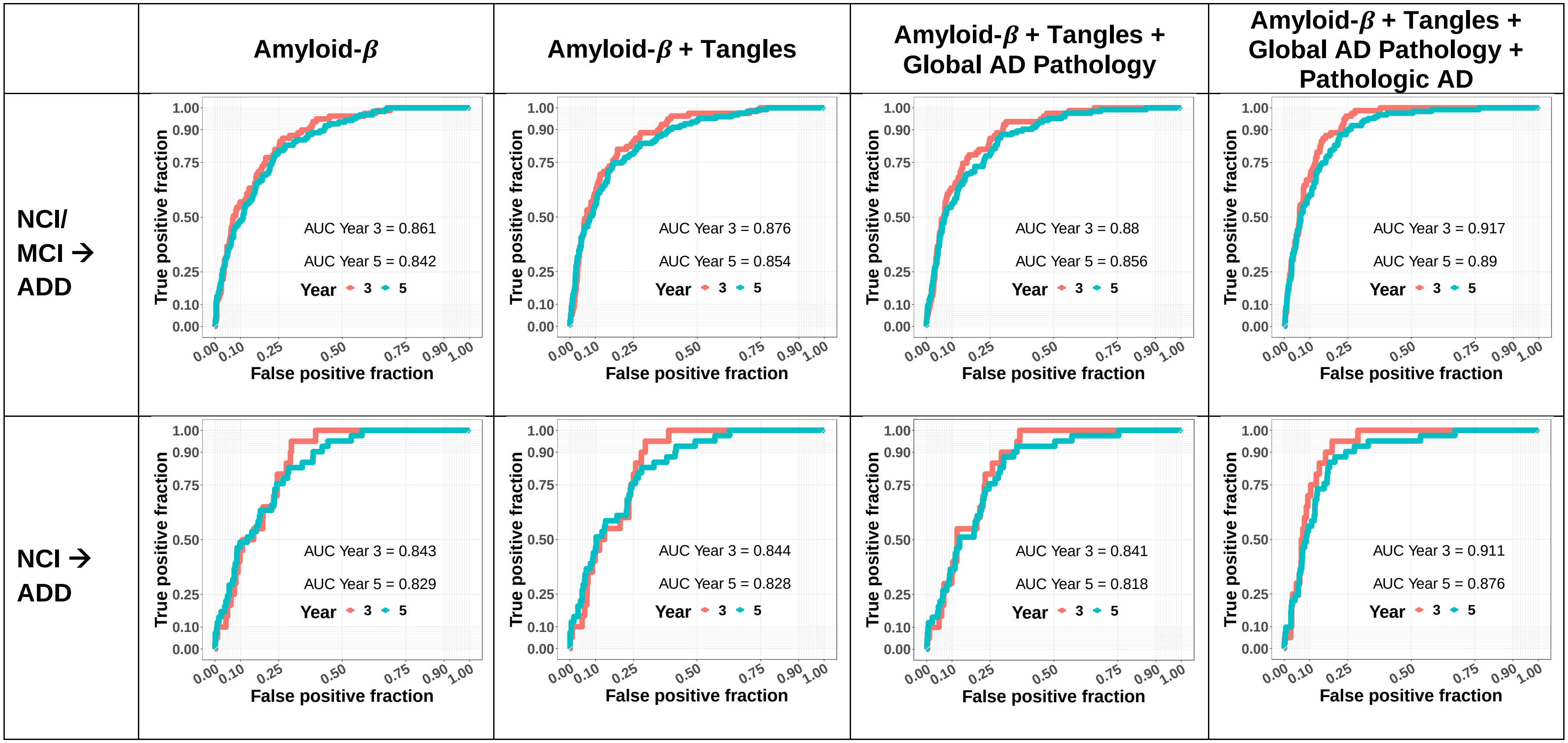
